# An optimised protocol for detection of SARS-CoV-2 in stool

**DOI:** 10.1101/2021.01.11.20248606

**Authors:** T Li, E Garcia-Gutierrez, J Scadden, J Davies, C Hutchins, A Aydin, S Romano, L Sayavedra, J O’Grady, A Narbad

## Abstract

**Aim:** SARS-CoV-2 has been detected in stool samples of COVID-19 patients, with potential implications for faecal-oral transmission. Compared to swab samples, the complexity of the stool matrix poses a challenge in the detection of the virus that has not yet been solved. The aim of this study was to establish a sensitive and reliable method for detecting SARS-CoV-2 in stool samples.

**Methods:** Stool samples from individuals free of SARS-CoV-2 were homogenised in saline buffer and spiked with a known titre of inactivated virus ranging from 50 to 750 viral particles per 100 mg stool. Debris was removed via centrifugation and supernatants were concentrated by ultrafiltration. RNA was then extracted from the concentrated material using a commercial kit and SARS-CoV-2 was detected via real-time reverse-transcription polymerase chain reaction (RT-qPCR) using the CDC primers and probes.

**Results:** The RNA extraction procedure we used allowed the detection of SARS-CoV-2 via RT-qPCR in most of the stool samples tested. We could detect as few as 50 viral particles per 100 mg of stool. However, high variability was observed across samples at low viral titres. The primer set targeting the N1 region provided more reliable and precise results and for this primer set our method had a limit of detection of 1 viral particle per mg of stool.

**Conclusions:** Here we describe a sensitive method for detecting SARS-CoV-2 in stool samples. This method can be used to establish the persistence of SARS-CoV-2 in stool and ensure the safety of clinical practices such as faecal microbiota transplant (FMT).

## 1. Introduction

The COVID-19 global pandemic, caused by SARS-CoV-2, poses an imminent threat to the global population. From December 2019 until the 16^th^ of December 2020, the number of confirmed cases stands at 72.7 million and rising, leading to an unprecedented challenge on health systems internationally. SARS-CoV-2 causes severe acute respiratory syndrome - infecting human cells by binding to the receptor angiotensin converting enzyme 2 (ACE2). ACE2 is an inflammation regulator on the epithelial cells of the lung, liver, and gastrointestinal tract ^1,2^. It has been reported that gastrointestinal symptoms, such as diarrhoea, nausea, and vomiting, may be observed in up to 61% of cases ^3^. These gastrointestinal symptoms may be linked to the severity of the COVID-19 disease based on viral load and degree of viral replication in the gut ^3–6^. SARS-CoV-2 RNA has been detected in the stool during infection, as well as when the patients have apparently recovered, and the nasal swab is negative ^7^. Viable SARS-CoV-2 has been isolated from stool samples ^7–9^, which suggests that there is a potential risk of faecal-oral transmission ^10–14^. Hence, monitoring the viral load in stool is of crucial importance to maintain public health and limit viral spreading. A few studies have reported that the viral load in stool samples (10^2^-10^7^ genome copies mL^-1^) is several orders of magnitude lower than in saliva (10^8^ genome copies mL^-1^) ^15,16^. However, methods for the detection of the virus in stool have been poorly described. Robust and reliable methods are an urgent need, as microbiota-based therapies such as faecal microbiota transplantation (FMT) would need to rely heavily on the accurate screening of donor stools to ensure the absence of SARS-CoV-2 and guarantee patient safety ^17^.

For nasal swabs and saliva samples, RT-qPCR is the most used diagnostic tool for detecting SARS-CoV-2, with many assays targeting the SARS-CoV-2 nucleocapsid (N) gene ^18,19^. The commonly used Center for Disease Control and Prevention (CDC) test targets two regions of the N gene (N1 and N2). However, the faecal matrix has properties distinct from those of respiratory samples ^20–22^ and this makes the reliable detection of SARS-CoV-2 challenging. Recently, a few methods have been described for the detection of SARS-CoV2 in stool samples ^17,23,24^. However, the potentailly low concentration of SARS-CoV2 in faeces and the unique features of the sample matrix require optimized protocols to improve the recovery of viral RNA and increase our ability to detect the virus in stool samples. To address this need, we developed a reliable and sensitive method for SARS-CoV-2 detection in stool.

## 2. Methods

### 2.1 Sample preparation and RNA extraction

Stool samples collected either before the COVID-19 outbreak (October 2019), or from donors who did not display and had not displayed symptoms of COVID-19 were weighed (100 mg) and homogenised in saline solution (0.89% w:v NaCl) with a ratio of 1:10 (w:v; 100 mg in 1 mL) by vortexing for at least 1 minute. In an initial screening phase, we assessed the effect of an ultrafiltration step aimed at enriching viral particles (vp) before RNA extraction using a single stool sample. For these tests, we spiked the homogenised stool with different volumes of AMPLIRUN® TOTAL SARS-CoV-2 CONTROL (VirCell Microbiologists, Spain) and then either directly extracted RNA using the QIAamp Viral RNA Mini Kit (Qiagen, UK; CN 52906) or first enriched the viral particles by ultrafiltration and then extracted the RNA, as described below. Since the ultrafiltration step improved SARS-CoV-2 detection we added this step in all further RNA extractions.

All the following experiments were performed by spiking the homogenised stool samples with the inactivated stock solution of SARS-CoV-2 positive Q control, SCV2QC01-B, Qnostics (UK). We used this stock because it has been precisely quantified by the manufacturing company using digital PCR. The stock is available by the supplier (Randox Biosciences, UK) in transport media at a viral concentration of 10,000 digital copies (dC) mL^-1^. The homogenised stool samples (1 mL) were spiked with different concentrations of viral particles, centrifuged at 4,000 g for 10 minutes, and then supernatants were filtered through 0.22 µM syringe filters. Virus enrichments were then performed using ultrafiltration tubes (Sigma, UK; CN UFC810024) by loading 500 µl of the filtrate and centrifuging at 2,500 g for 10 minutes. The concentrated samples (around 50 µl) were used for RNA extraction using QIAamp Viral RNA Mini Kit, following the protocol provided by the manufacturer. Finally, the viral RNA was eluted in two aliquots of 40 µl buffer AVE supplied with the kit as recommended by the manufacturer to increase the yield. An overview of the method we developed is reported in Fig 1A. We first used three stool samples to assess the lowest amount of virus detectable with our approach. The same samples were then used to assess the effect of the solution used to homogenise the stool (Table 1). This was done by repeating the above protocol and homogenising the stool not in saline solution but in AVL Buffer (Qiagen, UK; CN 19089), as recommended in the QIAamp Viral RNA Mini Kit manufacturer manual. 24 additional stool samples were then used to verify the variability of the assay across stool types and to infer the sensitivity and specificity of our protocol by extracting RNA after spiking them with 0, 100 and 200 vp per 100 mg. Finally, an additional stool sample was used to calculate the limit of detection (LoD) of our method. An overview of the experiments performed in this study is reported in Fig 1B.

**Table 1.** Ct values of RT-qPCR using RNA extracted from sample homogenised in saline buffer and AVL buffer

| Viral load<sup>a</sup> | Saline buffer |  |  |  |  |  | AVL buffer |  |  |  |  |  |
| --- | --- | --- | --- | --- | --- | --- | --- | --- | --- | --- | --- | --- |
|  | Stool 1 |  | Stool 2 |  | Stool 3 |  | Stool 1 |  | Stool 2 |  | Stool 3 |  |
|  | N1-mean | N2-mean | N1-mean | N2-mean | N1-mean | N2-mean | N1-mean | N2-mean | N1-mean | N2-mean | N1-mean | N2-mean |
| 0 | - | - | - | - | - | - | - | - | - | - | - | - |
| 50 | 34.8 | 36.2 | - | - | - | 33.5 | 35.2 | - | - | - | - | - |
| 100 | 34.8 | 36.3 | - | - | 34.2 | 32.5 | 34.9 | 35.2 | - | - | - | - |
| 150 | 34.6 | 35.3 | 35.8 | - | 34.9 | 33.9 | 33.8 | 34.7 | 36.4 | - | - | - |
| 200 | 34.2 | 34.0 | 35.9 | - | 34.2 | 33.0 | 33.5 | 33.9 | 35.8 | - | 37.2 | - |
| 250 | 34.0 | 34.6 | 35.4 | - | 34.8 | 33.6 | 35.0 | 36.0 | 36.7 | - | 36.0 | - |
| 500 | 32.8 | 33.6 | 34.2 | - | 33.7 | 33.5 | 35.1 | 36.3 | 36.2 | - | 35.4 | 36.7 |
| 750 | 32.7 | 33.7 | 35.4 | 33.8 | 33.5 | 33.9 | 36.0 | - | 36.6 | - | 33.1 | 34.8 |

<sup>a</sup>Numbers of viral particles spiked in 100 mg of stool - = Sample showed no Ct value

**Figure 1.**
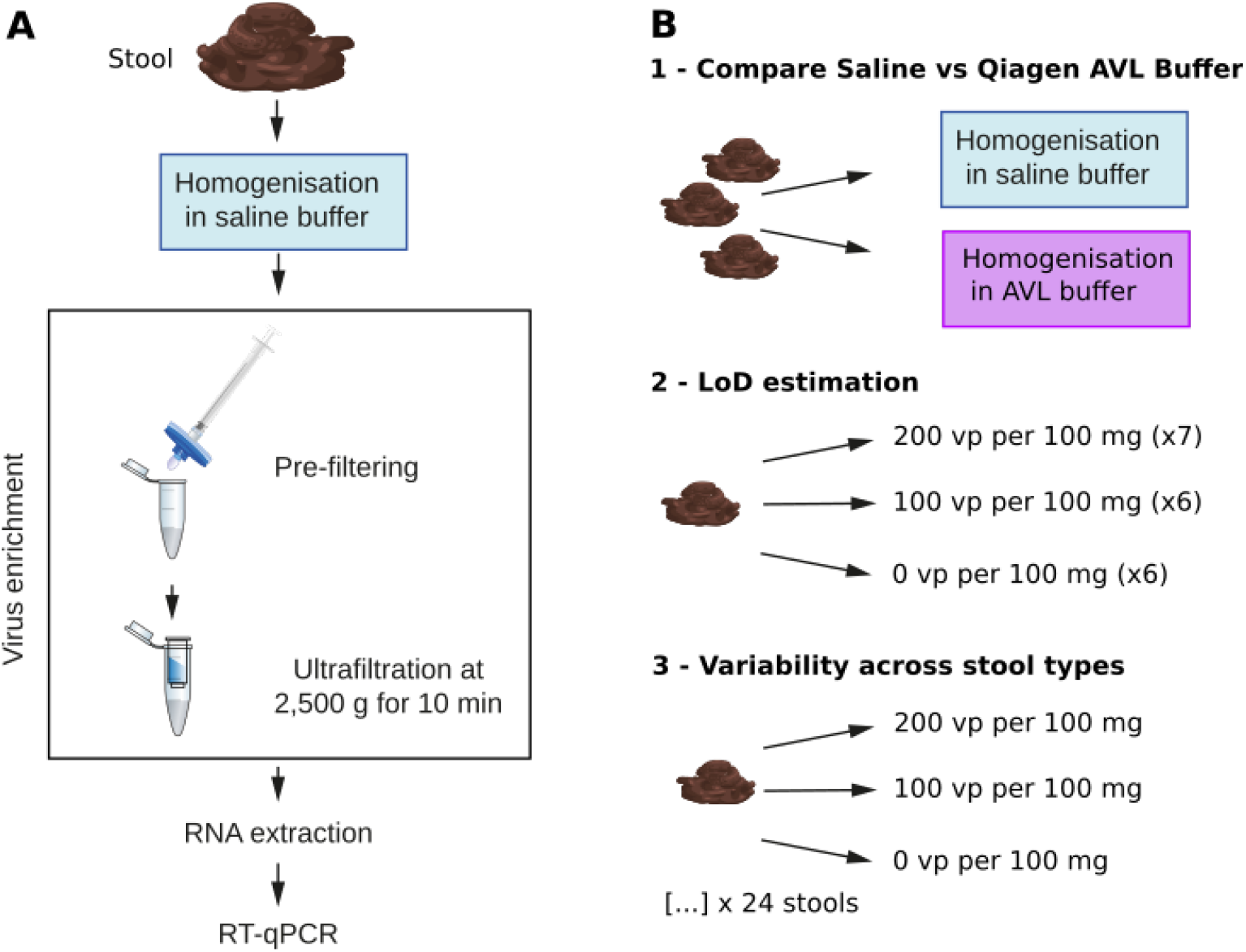
**A)** Overview of our optimised method for SARS-CoV-2 detection in stool samples. **B)** Schematic representation of the experiments conducted.

The LoD, defined as the lowest concentration of virus at which all samples had at least one positive RT-qPCR replicate, was estimated as follows. A freshly collected stool sample was diluted in saline solution in the ratio specified above and then stored at -20° C until further processing. After thawing, the homogenised stool sample was aliquoted and six and seven replicates were spiked with 100 and 200 vp per 100 mg, respectively. Six non-spiked replicates were also prepared. Samples were processed as described, and virus enrichment was performed using between 500 and 600 µl of supernatant. RNA was extracted as described. To verify the absence of contamination in the reagents, we also performed RNA extractions using kit reagents only. Finally, as positive control, we extracted RNA from 10 µL of the original stock used to spike the samples.

### 2.2 RT-qPCR assay

Primer sets N1 and N2 (Integrated DNA Technologies, Belgium, 10006713) were used for identifying SARS-CoV-2 with the Probe 1-Step Go No Rox or Probe 1-Step Go Rox kits (PCR Biosystems) ^25^. RT-qPCR was then performed in a Roche LightCycler® 480 Instrument II or a StepOnePlus™ Real-Time PCR System (Applied Biosystems™) using the following conditions: 50 ºC 10 min, 95 ºC for 2 min, 45 cycles of 95 ºC 5 s, 55 ºC 30 s, followed by 40 ºC 30 s. The Ct values were calculated and samples were considered as positive only if they showed Ct lower than 40 cycles. RT-qPCR was always performed using two technical replicates. As the aim of this study was to verify the variability in detecting SARS-CoV-2 in stool samples with the intention of reporting robust guidelines for screening FMT material, we considered positive also samples for which only 1 RT-qPCR technical replicate gave positive amplification. To determine the 95% confidence interval for the specificity and the sensitivity we used the *binom*.*test* function in the R software^26^.

## 3. Results

Several recent studies indicate that viable SARS-CoV-2 can be detected in stool samples of COVID-19 patients ^7,8^, suggesting that a possible risk for faecal-oral transmission exists. However, methods for the detection of SARS-CoV-2 in stool have been poorly assessed so far, and an optimized protocol is currently missing. Here, we describe an optimized protocol (Figure 1A) to improve the detection of SARS-CoV-2 in stool samples. We performed our experiments using samples collected either before the current pandemic started (October 2019) or from healthy donors who did not display and had not displayed symptoms of COVID-19. We used a commercially available SARS-CoV-2 stock, which was quantified using digital PCR by the manufacturer and was therefore used to infer the limit of detection of our approach. The extracted RNA samples were then used to detect SARS-CoV-2 via RT-qPCR with primer sets N1 and N2 using established protocols ^25^. We assessed how various steps throughout the RNA extraction influence the detection of SARS-CoV-2 in stool and report recommendations for optimizing these procedures in future clinical settings.

First, we spiked different volumes of an inactivated viral stock in stool samples and extracted the RNA with or without ultrafiltration (Supplementary Table 1). We obtained positive amplifications for both the N1 and N2 regions using both approaches. Without ultrafiltration we obtained positive amplifications with both N1 and N2 primer sets down to 2900 vp per 100 mg. In contrast, the addition of ultrafiltration allowed us to detect positive amplifications with both primer sets down to 725 vp per 100 mg (Supplementary Table 1). Hence, for all further tests we included an ultrafiltration step in our protocol. We then wanted to assess what viral concentration was more reliably detected across stool types. Thus, we used a viral stock that had been accurately quantified using digital PCR and we spiked three stool samples using concentrations ranging from 0 to 750 vp per 100 mg. We were able to detect SARS-CoV-2 in all stool samples we tested. The lowest concentration we could detect was 50 vp per 100 mg (Table 1), but a high variability amongst samples was observed for the lower concentrations (50-200 vp per 100 mg). This variability might be the result of the stool characteristics, for example, mucus and fibre content, as it has been previously reported that stool features can inhibit molecular assays ^20–22^.

We then selected the two lowest concentrations that gave reliable results (100 and 200 vp per 100 mg) to estimate the limit of detection (LoD) of our method. The stool sample used to determine the LoD was exclusively used for this experiment (i.e. was not used to create the previous datasets). We performed repeated extractions after spiking the homogenized stool sample with 100 vp per 100 mg (6 replicates in total) and 200 vp per 100 mg (7 replicates in total). As negative control, we also extracted 6 non-spiked stool aliquotes all of which resulted negative in the RT-qPCR assay. For the N1 primer set all samples spiked with either 100 or 200 vp per 100 mg resulted positive in the RT-qPCR assay (Table 2). For the N2 primer set instead, 3 out of 6 and 7 out of 7 samples gave positive amplification for the 100 and 200 vp per 100 mg spikes, respectively (Table 2). Based on this data our approach has a LoD, defined as the lowest concentration at which all tested samples gave positive results in at least one RT-qPCR replicate, of 100 vp per 100 mg (1 vp mg^-1^) for the N1 primer set and 200 vp per 100 mg (2 vp mg^-1^) for the N2 primer set. These data are consistent with previous reports that highlighted the higher sensitivity of the N1 assay compared to the N2 assay ^27^. This LoD corresponds to 1000 vp g^-1^ for N1 and 2000 vp g^-1^ for N2 and to the best of our knowledge is the lowest LoD so far described ^17,24^. It is worth noting that for practical reasons we took roughly half of the initial faecal slurry for RNA extraction (500-600 μL of the 1 mL in which 100 mg stool was homogenised). Hence, the Ct values we obtained are from roughly half the initial number of viral particles spiked, if we assume an homogeneous suspension. Moreover, following the recommendation in the RNA extraction kit manufacture’s protocol we eluted the extract twice with 40 μL buffer. Although the elution with this volume (2 × 40 μL) might increase the total yield of RNA, an elution with lower volume could result in a more concentrated extract and therefore, increase the sensitivity of the RT-qPCR assay. Therefore, we believe that our method has room for further improvements by enhancing the separation between debris and supernatant to recover higher fractions of the slurry used in the RNA extraction and by using smaller volumes for RNA elution.

**Table 2.** Overview of the limit of detected (LoD) experiments

|  | N1 |  | N2 |  |
| --- | --- | --- | --- | --- |
| Viral particles in 100 mg | 100 | 200 | 100 | 200 |
| Number of replicates | 6 | 7 | 6 | 7 |
| SARS-CoV-2 positive<sup>a</sup> | 6 (100%) | 7 (100%) | 3 (50%) | 7 (100%) |
| Mean Ct (st. dev) | 32.6 (±0.8) | 31.4 (±0.5) | 32.8 (±0.4) | 32.1 (±0.5) |

<sup>a</sup> Samples were considered positive if at least one RT-qPCR replicate showed Ct values < 40.

Finally, we tested our method on 24 additional stool samples to estimate the variability in detection deriving by stool type and infer the specificity and sensitivity of our assay. We were able to detect the virus reproducibly and consistently across the majority of the samples with average Ct values ranging from 32.6 to 38.2 and from 31.4 to 37.9 for the N1 and N2 regions, respectively (Figure 2). Using these 24 samples we estimated that our method has a sensitivity of 100% for both the N1 and N2 primer set, and a specificity of 96% and 92% for the N1 and N2 primer sets, respectively (Table 3). The false-positive results might derive from contamination that occurred during sample preparation or could be a consequence of the virus enrichment in our protocol. Stool samples contain a high diversity of mostly uncharacterized viruses ^28^, and RNA viruses are among the lesser known viruses within the human virome ^29^. Hence, by enriching viral particles we increase the chances of extracting RNA from novel viruses that might be unspecifically amplified in the RT-qPCR assay, leading to false-positive results. Additional tests are needed to verify this hypothesis.

**Table 3.** Sensitivity and specificity of the protocol we developed. Data refer to the 24 stool samples spiked with 100 viral particles per 100 mg. The 95% confidence interval is reported in brackets. A sample is considered positive if a single RT-qPCR replicate with Ct < 40 is detected for either the N1 or N2 primer set.

| Regions of N gene amplified | Sensitivity (N = 24) | Specificity (N = 24) |
| --- | --- | --- |
| N1 | 24/24 (100%, CI = 86-100) | 23/24 (96%, CI = 79-100) |
| N2 | 24/24 (100%, CI = 86-100) | 22/24 (92%, CI = 73-99) |

**Figure 2.**
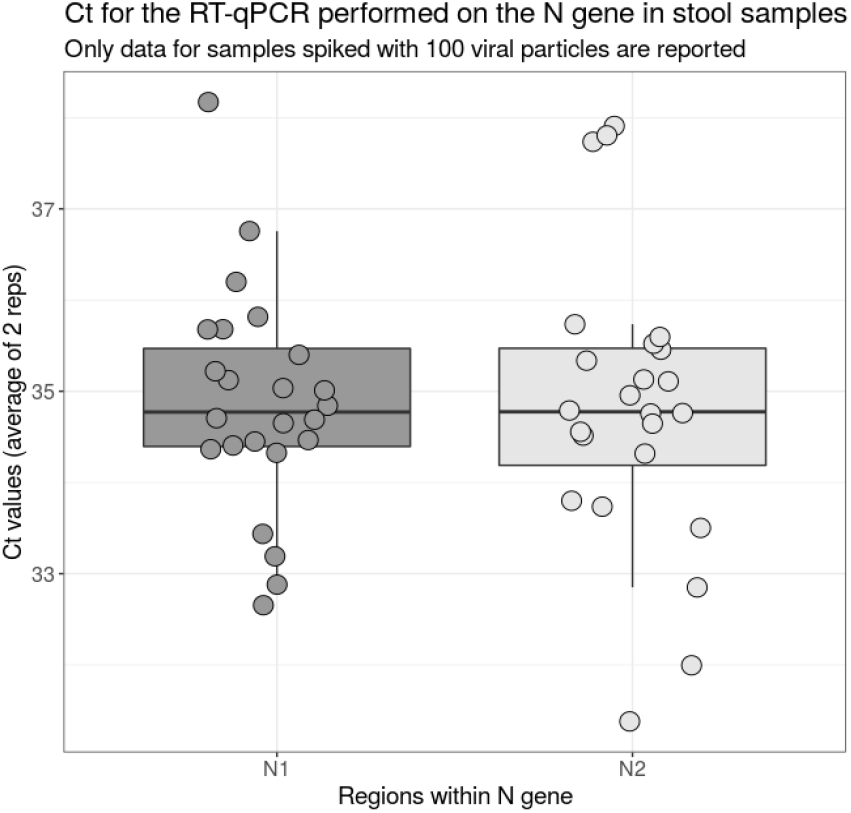
Ct values obtained for the N1 and N2 regions used to detect SARS-nCoV-2 in stool samples. Only data for the samples spiked with 100 vp per 100 mg are reported. In five samples only one replicate gave positive results, which have been included in the graphs.

We consistently detected viral particles in stool samples diluted in saline buffer. We then tested whether homogenising the faecal material in the Qiagen AVL lysis buffer instead of saline buffer might affect RNA recovery. For this additional test, we used the same three stool samples initially used to assess the lowest viral load detectable, but homogenised them in Qiagen AVL buffer. When we used the Qiagen AVL buffer a higher variability was observed across samples (Table 1). We observed that the faecal material does not fully homogenise in the lysis buffer (Supplementary Figure 1), suggesting that this might potentially affect the release of viral particles from the stool matrix. This observation is particularly relevant because in several studies RNA has been extracted from stool samples by applying the standard manufacturers’ protocol of the extraction kits ^14,30^. These are not necessarily optimized for stool and recommend dissolving samples in the extraction buffer (e.g. Qiagen AVL buffer). Our data indicate that depending on the stool matrix type, this procedure can reduce the efficiency of the RNA extraction and possibly underestimate the virus detection.

## 4. Conclusions

Here we assess the technical challenges encountered while screening stool material for the presence of SARS-CoV-2. We describe a robust approach to detect SARS-CoV-2 in stool samples, having an LoD of 1 and 2 vp mg^-1^ for the N1 and N2 primer set, respectively. Although we could detect as low as 50 vp per 100 mg, a high variability can be observed between stool sample types when viral concentrations are low. We demonstrated that following manufacturer’s RNA extraction protocols may not be sufficient to detect SARS-CoV-2 in stool samples as stool consistency and homogenisation media can affect the downstream assays. To improve detection, we suggest to homogenise the stool samples in saline solution first, then concentrate the viral particles with ultrafiltration. Our method is sufficiently reliable to monitor the prevalence and persistence of the viral particles in the gut and can help in determining the safety of samples intended for use in FMT applications. To ensure the maximal safety for the patients, we propose that FMT donors should be screened following the recommendations here, and donor stool should be excluded even if a single N region (N1 or N2) or a single replicate per region gives a positive result due to the variability introduced by the stool matrix.

## Data Availability

All data are reported in the submitted manuscript

## Acknowledgements

We thank Emma Flannagan and Dr David Vauzour for providing samples, Lee Kellingray, George Savva, and Sharlize Pedroza-Matute for fruitful discussions and Gemma Kay for provision of reagents. This work was supported by funding from the QIB, the Biotechnology and Biological Sciences Research Council (BBSRC) Impact Accelerator Account (BB/S506679/1), Institute Strategic Programme Gut Microbes and Health (BB/R012490/1) and its constituent projects BBS/E/F/000PR10355 and BBS/E/F/000PR10356, Institute Strategic Programme Microbes in the Food Chain BB/R012504/1 and its constituent projects BBS/E/F/000PR10348, BBS/E/F/000PR10349, BBS/E/F/000PR10351, and BBS/E/F/000PR10352.

## Ethics statement

Faecal samples were provided by different donors either via the NRP Biorepository from a study approved by the Ethical Review Committee of the University of East Anglia, Norwich (R204719) registered at http://www.clinicaltrials.gov/ (NCT03679533) or by a study approved by the QIB Human Research Governance committee (IFR01/2015) and registered at http://www.clinicaltrials.gov/ (NCT02653001).

## Supplementary materials

**Supplementary Figure 1.**
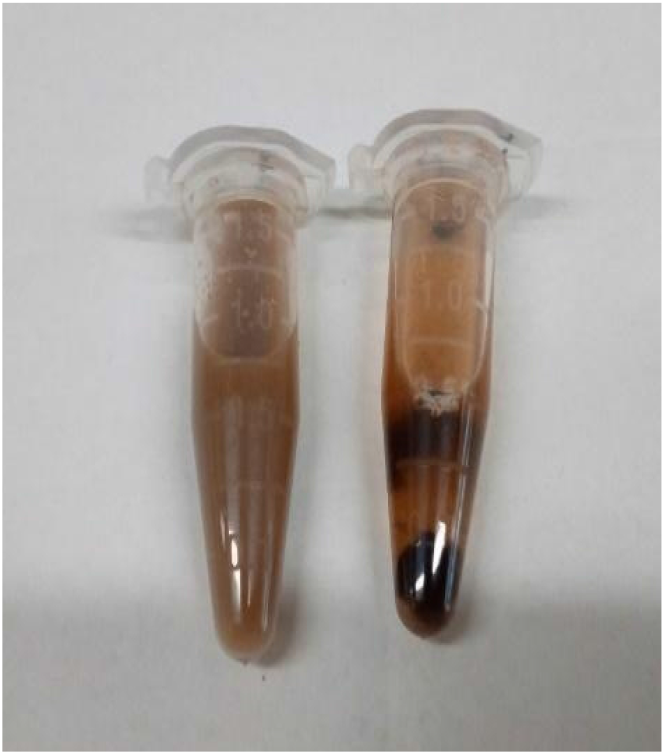
Faecal samples mixed and resuspended in 1 mL of saline buffer (left) resulted in a better homogenisation of the stool as compared to using AVL buffer (right).

**Supplementary Table 1.** Ct values obtained from stool samples spiked with different amounts of viral particles used to extract RNA with or without ultrafiltration.

| Spiked viral particles (vp in 100 mg) | Without ultrafiltration |  |  |  | With ultrafiltration |  |  |  |
| --- | --- | --- | --- | --- | --- | --- | --- | --- |
|  | N1 | N1 | N2 | N2 | N1 | N1 | N2 | N2 |
| 2900 | 32.3 | 31.9 | 31.4 | 31.3 | nt | nt | nt | nt |
| 1450 | 32.6 | 33.6 | - | - | 32.7 | 32.6 | 31.3 | 31.6 |
| 725 | nt | nt | nt | nt | 33.2 | 32.5 | 33.5 | 33.6 |

nt = not tested
- = Sample showed no Ct value

